# COMPENSATED HYPOGONADISM IN MEN WITH SICKLE CELL DISEASE

**DOI:** 10.1101/2020.04.21.20074666

**Authors:** Anna Paloma Martins Rocha Ribeiro, Carolina Santos Silva, Jean Carlos Zambrano, Juliana de Oliveira Freitas Miranda, Carlos Augusto Fernandes Molina, Cristiano Mendes Gomes, Eduardo de Paula Miranda, José de Bessa Júnior

## Abstract

**Introduction:** Sickle cell disease (SCD) is associated with the development of hypogonadism, but there is still controversy regarding its etiology and clinical implications.

**Objective:** To evaluate the prevalence of hypogonadism in a population of men with SCD and characterize its etiology.

**Methods:** We performed a cross-sectional study of 34 men with SCD aged > 18 years. Sociodemographic and clinical data, including anthropometric measurements (weight, height, and BMI), were obtained. Early morning blood samples were collected and total testosterone (TT), free testosterone (FT), luteinizing hormone (LH), follicle-stimulating hormone (FSH), a complete blood count, and hemoglobin electrophoresis were measured.

**Results:** Median age was 33 [26-41] years, and SS genotype was the most frequent (73.5%). The prevalence of eugonadism, compensated, and secondary hypogonadism was 67.5%, 26.4%, and 5.88%, respectively. No men with primary hypogonadism were identified in our sample. Those with compensated hypogonadism had also higher FSH levels than individuals with eugonadism; p < 0.001).

**Conclusion:** In our study population of men with SCD a high prevalence of compensated hypogonadism was identified, which is a controversial and distinct clinical entity that warrants monitoring and further research.

## INTRODUCTION

Sickle cell disease (SCD) is a relatively common genetic disease and comprises a group of disorders characterized by the presence of at least one hemoglobin S (1). The sickle gene mutation is common in sub-Saharan Africa and other parts of the world and it is estimated that more than 300 000 children are born each year with SCD, with millions of people currently affected across the globe (2,3), about two-thirds of them in Africa (ref). SCD has been recognized as a public health issue and a neglected problem by several key agencies, including the United Nations (UN) and the World Health Organization (WHO)(4). The chronic morbidity associated with SCD may lead to an increased socioeconomic burden and requires long-term quality of care (5) SCD is commonly associated with the development of hypogonadism, but there is controversy regarding its etiology, mechanisms, and clinical implications (5).

Hypogonadism is characterized by impaired testicular function, which may affect spermatogenesis and/or testosterone synthesis. Usually the diagnosis of hypogonadism requires identification of low serum testosterone (T) levels. Individuals with low T levels may be asymptomatic or present with a worse metabolic status, reduced energy, diminished physical performance, fatigue, depression, reduced motivation, poor concentration, infertility, reduced sex drive, erectile dysfunction and increased rates of all-cause mortality (6–8).

Although the development of androgen deficiency is currently considered multifactorial, male hypogonadism has been classically classified as hypergonadotropic (primary) or hypogonadotropic (secondary) according to its etiology (9). More recently a new clinical variant of hypogonadism defined as defined as compensated hypogonadism has been proposed, which is characterized by normal T and elevated LH levels. These authors suggested that compensated hypogonadism represents a distinct clinical state, which warrants monitoring despite its unknown etiology and consequences (8). Compensated hypogonadism has also been reported in 3% of patients with male infertility, with similar outcomes of those with primary hypogonadism (9).

Multiple theories have been proposed in attempts to explain the hypogonadism related to SCD. Zinc deficiency, socioeconomic factors, constitutional variables, and repetitive vaso-occlusive episodes in the testes and pituitary, are possible explanations though the definite cause remains unknown. Vaso-occlusion is the most widely accepted theory as recurrent microinfarctions in patients with SCD are commonly found in other organs and systems (10). Few studies have been designed to evaluate hormonal abnormalities in men with SCD and a more precise understanding is yet to be accomplished (11).

As studies evaluating detailed hormonal profiles in men with SCD are lacking, the aim of this study was to estimate the prevalence of hypogonadism in men with SCD and to characterize its etiology using a more comprehensive classification.

## MATERIALS AND METHODS

### Study design and population

This was a cross-sectional study involving 34 men with SCD aged 18 or older who were followed up at a local SCD referral center between January and December 2019. Men with a history of cryptorchidism, testicular tumors, testicular or inguinal surgery, or with acute onset of testicular at the time of the interview were excluded. A structured questionnaire, including sociodemographic (age, sex, self-reported race/color) and clinical variables (anthropometric measurements, type of hemoglobinopathy, therapy with hydroxyurea or NSAIDs, and the occurrence of priapism), was applied to participants. Anthropometric data (weight and height) were measured and used to calculate the BMI as weight in kilograms divided by height in meters squared (kg/m^2^).

### Hormonal evaluation

Blood samples were collected between 7 and 9 a.m. for the determination of total testosterone (TT), free testosterone (FT), LH, and FSH levels, as well as a complete blood count and hemoglobin electrophoresis. The methods used for the laboratory tests included: hydrodynamic focusing, flow cytometry, SLS-hemoglobin, and Giemsa microscopy for the complete blood count; electrochemiluminescence assays (Atellica IM® analyzer, Siemens Healthcare Diagnostics Inc., Tarrytown, NY, USA) for the determination of T, LH, and FSH levels; an equation involving TT, sex hormone-binding globulin (SHBG), and the association constant of albumin for T, assuming a fixed albumin concentration of 4.3 g/dL, for the calculation of FT(12); and high-performance liquid chromatography (HPLC) and capillary electrophoresis (Capillarys Hemoglobine) for the quantitation of hemoglobin fractions.

Subjects were classified into four groups according to T and LH levels(13): men were considered to have eugonadism if T ≥ 300 ng/dL and LH ≤ 9.4 mUI/mL; primary hypogonadism was defined as T < 300 ng/dL and LH > 9.4 mUI/mL; secondary hypogonadism as T < 300 ng/dL and LH ≤ 9.4 mUI/mL; and compensated hypogonadism as T ≥ 300 ng/dL and LH > 9.4 mUI/mL. FSH levels above 7.8 ng/dL were considered elevated (14).

All subjects were verbally and individually approached and provided informed consent. This study was approved by our Research Ethics

### Statistical analysis

Quantitative variables were presented as medians and interquartile ranges, while nominal variables were expressed as absolute values, percentages, or fractions. The Mann-Whitney U test was used to compare continuous variables, while the Fisher’s test was used to compare categorical variables. A p < 0.05 was considered statistically significant, and 95% confidence intervals were presented as a measure of precision. GraphPad Prism, version 8.0.3, San Diego-CA, USA, was used for data analysis.

## RESULTS

Our sociodemographic data are detailed in **Table 1**. We assessed 34 men with a median age of 33 years [26-41], most of whom had an SS genotype (73.5%) and were black or brown (94.1%). Five (14.7%) of them were on continuous hydroxyurea therapy and all had been medicated with NSAIDs, with ibuprofen being the most commonly used drug.

**Table 1.** Sociodemographic and clinical data of men with SCD.

| Variables | n (34) | % |
| --- | --- | --- |
| **SCD genotype** |  |  |
| Homozygous HbSS | 25 | 73.5 |
| Heterozygous SC/SB-thal | 9 | 26.5 |
| **Self-reported race/color** |  |  |
| Black | 21 | 61.8 |
| Brown | 11 | 32.4 |
| Yellow | 1 | 2.9 |
| Indigenous | 1 | 2.9 |
| **Hydroxyurea therapy** |  |  |
| No | 29 | 85.3 |
| Yes | 5 | 14.7 |
| **NSAIDs use** |  |  |
| No | 0 | 0 |
| Yes | 34 | 100 |
| **Education level** |  |  |
| Primary education | 7 | 20.6 |
| Secondary education | 20 | 58.8 |
| Higher education | 7 | 20.6 |
| **Occupation** |  |  |
| Active<sup>a</sup> | 6 | 17.7 |
| Inactive<sup>b</sup> | 28 | 82.3 |
| **History of priapism** |  |  |
| Yes | 18 | 52.9 |

Fifty percent of patients had LH levels of 5.92 mUI/mL [4.36 – 9.42]. Median FSH levels were 6.01 mUI/mL [4.08 – 8.96], clinical data are detailed in **Table 2**. In our sample, no men were diagnosed with primary hypogonadism, whereas 23 (67.6%) were classified as eugonadal and 2 (5.8%) as having secondary hypogonadism. In addition, compensated hypogonadism was identified in 9 men (26.4%) (**Figure 1**).

**Table 2.** Anthropometric and laboratory data of men with SCD

| Variables | Median [p25-p75]* |
| --- | --- |
| Weight (kg) | 61.5 [56.3 – 67.3] |
| Height (m) | 1.7 [1.64 – 1.75] |
| BMI (kg/m<sup>2</sup>) | 21.6 [19.2 – 23.8] |
| Hemoglobin (g/dL) | 9.75 [8.05 – 12.0] |
| Total testosterone (ng/dL) | 582.9 [428.8 – 685.9] |
| Free testosterone (ng/dL) | 9.85 [8.07 – 11.37] |
| LH (mUI/mL) | 5.92 [4.36 – 9.42] |
| FSH (mUI/mL) | 6.01 [4.08 – 8.96] |

\*Data are expressed as medians and interquartile ranges (25<sup>th</sup> and 75<sup>th</sup> percentiles)

**Table 3.**
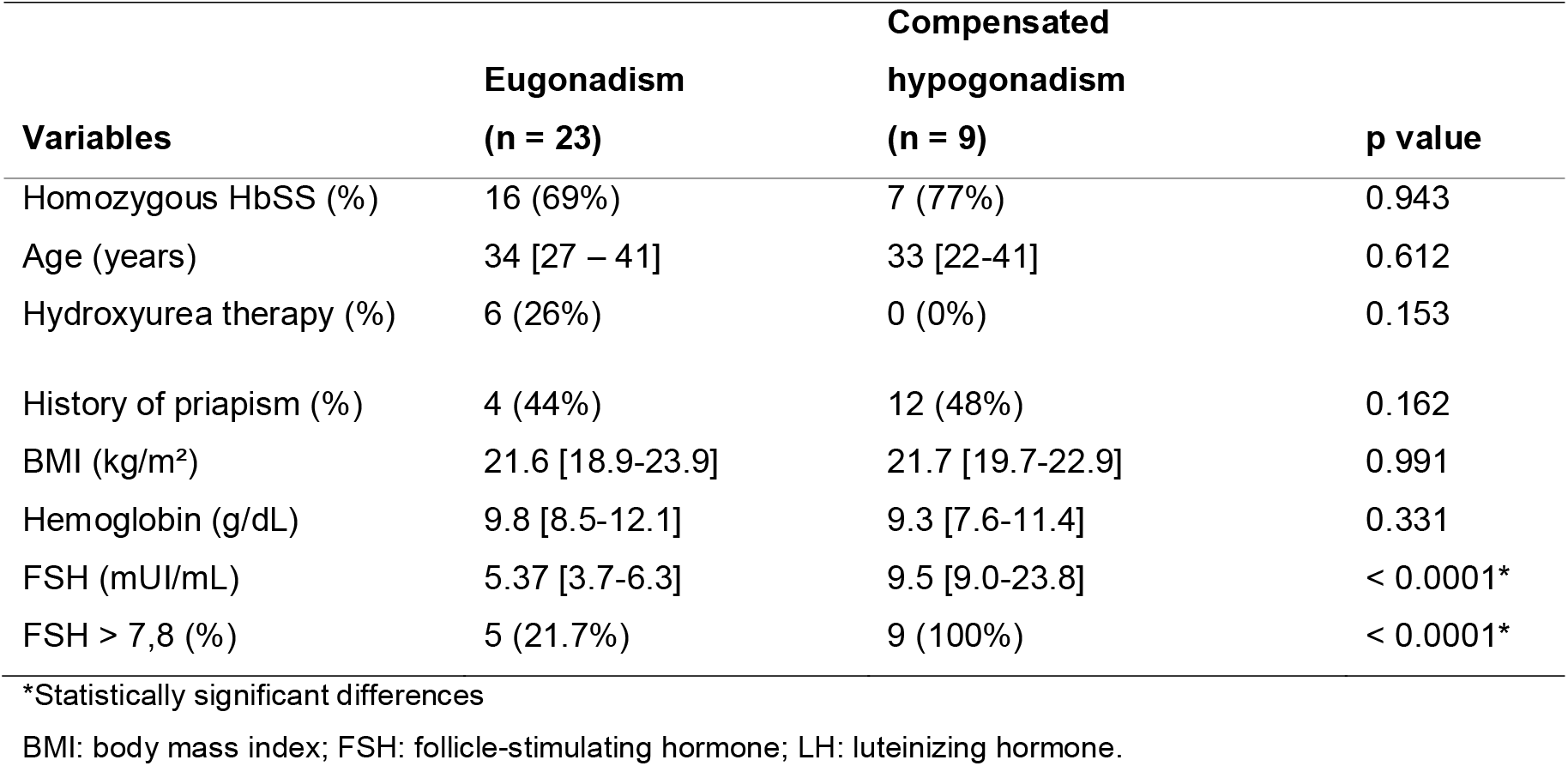
Clinical, anthropometric, and laboratory data of eugonadal men with SCD in comparison to those with compensated hypogonadism.

| **Variables** | **Eugonadism**<br>**(n = 23)** | **Compensated**<br>**hypogonadism**<br>**(n = 9)** | **p value** |
| --- | --- | --- | --- |
| Homozygous HbSS (%) | 16 (69%) | 7 (77%) | 0.943 |
| Age (years) | 34 [27 – 41] | 33 [22-41] | 0.612 |
| Hydroxyurea therapy (%) | 6 (26%) | 0 (0%) | 0.153 |
| History of priapism (%) | 4 (44%) | 12 (48%) | 0.162 |
| BMI (kg/m<sup>2</sup>) | 21.6 [18.9-23.9] | 21.7 [19.7-22.9] | 0.991 |
| Hemoglobin (g/dL) | 9.8 [8.5-12.1] | 9.3 [7.6-11.4] | 0.331 |
| FSH (mUI/mL) | 5.37 [3.7-6.3] | 9.5 [9.0-23.8] | < 0.0001* |
| FSH > 7,8 (%) | 5 (21.7%) | 9 (100%) | < 0.0001* |

\*Statistically significant differences
BMI: body mass index; FSH: follicle-stimulating hormone; LH: luteinizing hormone.

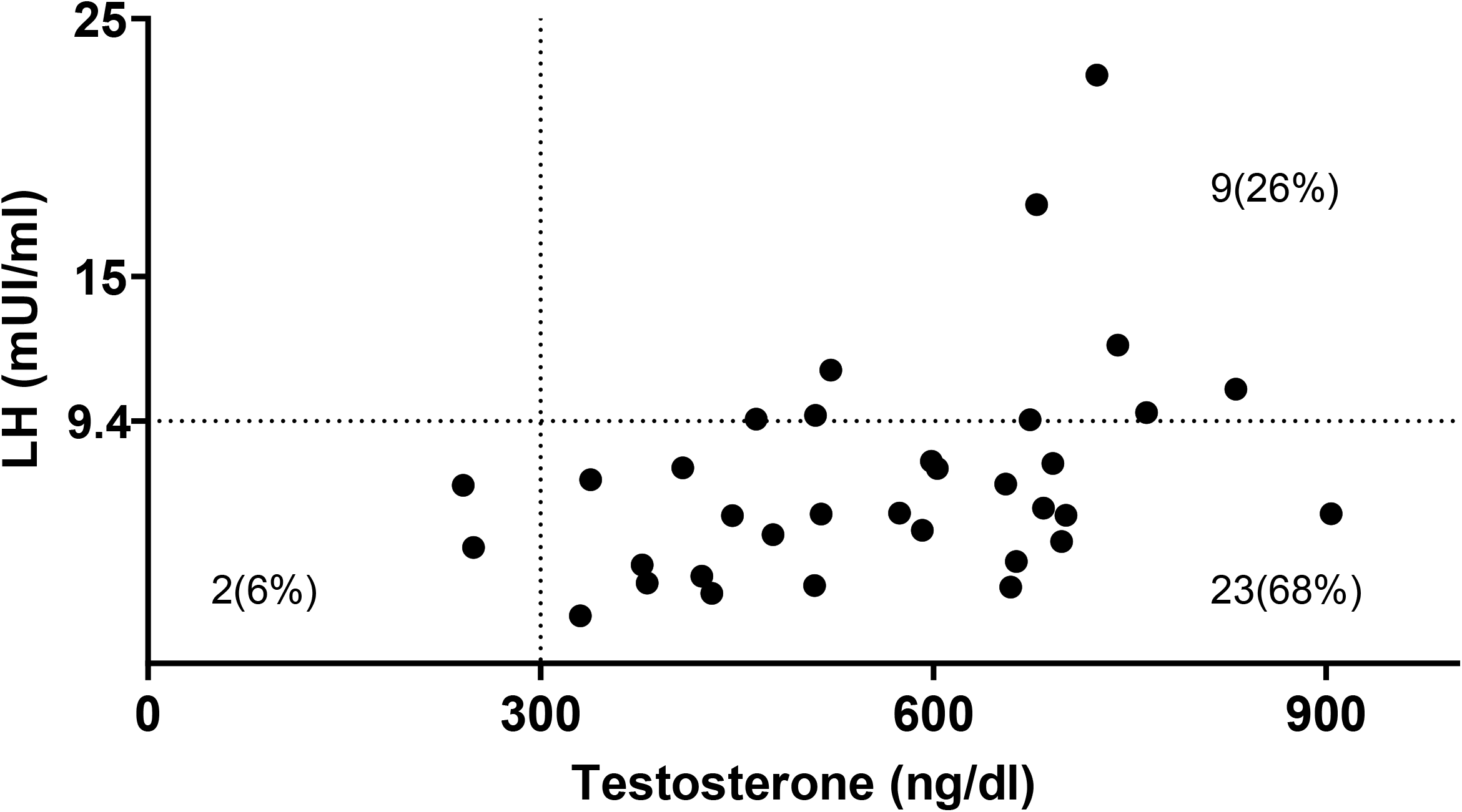

We found no differences regarding SCD genotype, anthropometric measurements, or disease severity between eugonadal men and those with compensated hypogonadism. Median FSH levels among men with compensated hypogonadism were significantly higher than among eugonadal men. The proportion of men with FSH levels above 7.8 mUI/mL was significantly higher among those with compensated hypogonadism (p < 0.0001) (**Table 2**).

## DISCUSSION

Gonadal dysfunction, disturbances of the hypothalamic-pituitary-testicular axis, and the etiology of hypogonadism in patients with SCD are controversial issues, and there are still many uncertainties about the pathophysiology of these conditions and their clinical significance(11). We investigated the prevalence of different types of hypogonadism in men with SCD according to the classification described by Tajar et al (2010) in the EMAS. The prevalence of compensated hypogonadism in our sample of men with SCD was 26.4%, a figure considerably higher than those reported in similar studies, 9.5%(13) and 3%(14), respectively.

Tajar at al. (2010) were the first to explore the concept of compensated hypogonadism in a study with 3369 community-dwelling men aged 40-79 years from eight European centers. They also introduced the concept that different hypogonadism categories may present with different clinical features. According to this study, sexual complaints were more frequently reported in cases of primary hypogonadism, whereas isolated physical symptoms such as inability to walk long distances or perform brisk physical activity were more common in compensated hypogonadism (8).

A retrospective study involving 4173 men with sexual dysfunction by Corona et al., reported that 4.1% of individuals had compensated hypogonadism and were more likely to present with both increased frequency of psychological symptoms, such as anxiety, obsessive-compulsive symptoms, and depression; and increased cardiovascular mortality in comparison to those with primary or secondary hypogonadism. The authors hypothesized that compensated hypogonadism could be a surrogate marker of an underlying disease, rather than a new clinical entity(15).

Compensated hypogonadism also appears to be relevant in the elderly. Ucak et al. (2013) conducted a study with 250 men over 70 years of age with compensated hypogonadism and found their T and LH levels to be independently associated with worsening performance of activities of daily living, as well as deteriorating cognitive function, nutritional status, and mood, when compared to healthy controls. The authors concluded androgen and LH levels should be assessed in elderly men, and those diagnosed with compensated hypogonadism should then undergo a physical and neuropsychiatric evaluation (16).

The clinical significance of compensated hypogonadism is still poorly understood, but this condition is known to be associated with both aging and an increased frequency of physical symptoms related to testosterone deficiency, but not with sexual complaints, as previously mentioned. These observations and its striking prevalence of 9.5% as reported in the EMAS raise the question of whether this subtype of hypogonadism warrants treatment. In, especially older men, may benefit from the inclusion of LH levels in the initial screening for hypogonadism (17).

Studies about compensated hypogonadism and its etiology are controversial and scarce in patients with SCD. Rhodes et al (2009)(18) did not found a statistically significant difference in the testosterone levels of 19 boys with SCD when compared to controls and so they were unable to detect primary hypogonadism. Özen et al. (2013)(19) studied 50 Turkish children aged 4 to 18 years and found, among the 35 boys included in the sample, one with hypergonadotropic (primary) hypogonadism and 3 with small testes and low testosterone, but with normal luteinizing hormone (LH) levels, suggesting this condition can be either primary or secondary. Abbasi and colleagues (1976)(20) analyzed hormone levels of 14 patients and found elevated concentrations of LH and follicle-stimulating hormone (FSH), as well as low testosterone (T), suggesting primary hypogonadism. Dada and Nduka (1980)(22) demonstrated reduced LH, FSH, and testosterone levels in 19 men with SCD, findings suggestive of hypothalamic axis dysfunction (secondary hypogonadism), rather than gonadal failure (primary hypogonadism). Martins et al(23) studied some aspects of compensated hypogonadism among 10 men with homozygous SCD. Despite the limited number of patients, the authors referred to compensated hypogonadism as a likely transient condition and considered it a potential state of androgen resistance(23).

All subjects with compensated hypogonadism in our sample had FSH levels above 7.8 mU/mL, a threshold previously reported as a predictor of impaired spermatogenesis(24). It is likely that the exocrine compartment might be impaired in while the endocrine compartment might still be enough to sustain appropriate T production. A cross-sectional study with 786 Caucasian-European discussed the concept of compensated hypogonadism among infertile subjects. The authors found that this condition had a similar clinical characteristic to those with primary hypogonadism, and both groups had the worst clinical outcomes in terms of impaired fertility. While this classification was not designed for the setting of male infertility, it could be useful in clinical practice to indicate impaired spermatogenesis(14).

It is important to highlight that all men in our sample had history of ibuprofen intake for acute pain episodes. Previous published data indicate a strong association between ibuprofen use and the elevation of LH levels, which might lead to compensated hypogonadism. In a randomized controlled trial including 31 men aged 18-35 years who received 600 mg of ibuprofen twice daily for 6 weeks, the authors showed ibuprofen use increased LH levels in 23% after 14 days and 33% after 44 days (p = 0.01). They also linked ibuprofen use with a reduction in anti-Müllerian hormone levels and postulated this drug would affect steroidogenesis by inhibiting the expression of related genes, thus resulting in HPT axis dysfunction (25). However, this study evaluated solely the impact of short-term course of ibuprofen on hormonal profile, and clinical significance of its effect in the long-term are probably negligible.

Although our cohort had only 34 participants, it is one of the largest studies in the scientific literature to evaluate hormonal profiles in men with SCD. We also identified a high prevalence of an underreported and poorly understood condition, which might be of clinical significance for monitoring and counseling patients. The main limitation of this study is perhaps its cross-sectional design, as no definitive explanation for this condition may be given. In addition, this sample came from a specialized center for patients with SCD and may not represent the general population of SCD.

## CONCLUSION

A high prevalence of compensated hypogonadism was identified in our sample of men with SCD, which may reflect early testicular injury and may progress to permanent dysfunction. It seems to be more prevalent in young men with SCD than in the general population of older men. Vaso-occlusive phenomena, testicular microinfarctions, changes in the testicular microenvironment, long-term or recurrent ibuprofen use, and genetic aspects might be associated with such findings. Although this clinical entity is not completely understood, continuous monitoring might be useful in preventing or anticipating additional clinical deterioration.

## Data Availability

Data are attached as supplement in submission to Plos One

## Disclosure statement

The authors report no conflicts of interest.

## Funding

This study received no external funding.

## Academic affiliation

This paper is part of the master’s dissertation of Anna Paloma Martins Rocha Ribeiro, Postgraduate Program in Public Health, State University of Feira de Santana.

## Notes

### Competing Interest Statement

The authors have declared no competing interest.

### Funding Statement

No external fundings

## REFERENCES

1. Bunn HF. Pathogenesis and treatment of sickle cell disease. N Engl J Med. 1997;337(9):762–9.

2. Wastnedge E, Patel S, Goh MY, Rudan I. The global burden of sickle cell disease in children under five years of age: a systematic review and meta-analysis. J Glob Health. 2018;8(2):1–9.

3. Piel FB, Hay SI, Gupta S, Weatherall DJ, Williams TN. Global Burden of Sickle Cell Anaemia in Children under Five, 2010 – 2050: Modelling Based on Demographics, Excess Mortality, and Interventions. PLoS One. 2013;10(7):1–14.

4. Mburu J, Odame I. Sickle cell disease: reducing the global disease burden. Int J Lab Hematol. 2019;41(February):82–8.

5. Pleasants S. A moving target: migration has led to an increase in the occurrence of sickle-cell disease in countries with previously low incidence of the disorder. Nature. 2014;515(11):1–3.

6. Mulhall JP, Trost LW, Brannigan RE, Kurtz EG, Bruce J, Chiles KA, et al. Evaluation and Management of Testosterone Deficiency: AUA Guideline. J Urol [Internet]. 2018; Available from: https://doi.org/10.1016/j.juro.2018.03.115

7. Livingston M, Kalansooriya A, Hartland AJ, Ramachandran S, Heald A. Serum testosterone levels in male hypogonadismLJ: Why and when to check — A review. Int J Clin Pract. 2017;(August):1–9.

8. Peterson MD, Belakovskiy A, Mcgrath R, Yarrow JF. Testosterone Deficiency, Weakness, and Multimorbidity in Men. Sci Rep [Internet]. 2018;8(5897):1–9. Available from: http://dx.doi.org/10.1038/s41598-018-24347-6

9. Jungwirth A, Diemer T, Kopa Z, Krausz C, Minhas S, Tournaye H. EAU Guidelines on Male Infertility. Vol. 1, European Association of Urology. 2018. 47 p.

10. Li M, Fogarty J, Whitney KD, Stone P. Repeated testicular infarction in a patient with sickle cell disease: A possible mechanism for testicular failure. Urology. 2003;62(3):551.

11. Huang AW, Muneyyirci-Delale O. Reproductive endocrine issues in men with sickle cell anemia. Andrology. 2017;5(4):679–90.

12. Vermeulen A, Verdonck L, Kaufman JM. A critical evaluation of simple methodsfor the estimation of free testosterone in serum. J Clin Endocrinol Metab. 1999;84(10):3666–72.

13. Tajar A, Forti G, O’Neill TW, Lee DM, Silman AJ, Finn JD, et al. Characteristics of secondary, primary, and compensated hypogonadism in aging men: Evidence from the European male ageing study. J Clin Endocrinol Metab. 2010;95(4):1810–8.

14. Ventimiglia E, Ippolito S, Capogrosso P, Pederzoli F, Cazzaniga W, Boeri L, et al. Primary, secondary and compensated hypogonadism: a novel risk stratification for infertile men. Andrology. 2017;5(3):505–10.

15. Corona G, Maseroli E, Rastrelli G, Sforza A, Forti G, Mannucci E, et al. Characteristics of compensated hypogonadism in patients with sexual dysfunction. J Sex Med. 2014;11(7):1823–34.

16. Ucak S, Basat O, Karatemiz G. Functional and nutritional state in elderly men with compensated hypogonadism. J Am Med Dir Assoc. 2013;14(6):433–6.

17. Lee JA, Kuchakulla M, Arora H, Kulandavelu S, Gonzalez E, Masterson TA, et al. Age induced nitroso-redox imbalance leads to subclinical hypogonadism in Male mice. Front Endocrinol (Lausanne). 2019;10(MAR):1–6.

18. Rhodes M, Akohoue SA, Shankar SM, Fleming I, An A, Yu C, et al. Growth Patterns in Children with Sickle Cell Anemia during Puberty. Pediatr Blood Cancer. 2009;53(4):635–41.

19. Özen S, Ünal S, Erçetin N, Taşdelen B. Frequency and Risk Factors of Endocrine Complications in Turkish Children and Adolescents with Sickle Cell Anemia. Turk J Hematol. 2013;30:25–31.

20. Abbasi AA, Prasad AS, Ortega J, Congco E, Oberleas D. Gonadal Function Abnormalities in Sickle Cell Anemia Studies in Adult Male Patients. Ann Intern Med. 1976;85:601–5.

21. Dada OA, Nduka EU. Endocrine function and haemoglobinopathies: Relation between the sickle cell gene and circulating plasma levels of testosterone, luteinising hormone (LH) and follicle stimulating hormone (FSH) in adult males. Clin Chim Acta. 1980;105(2):269–73.

22. Dada O, Nduka E. Endocrine function and haemoglobinopathies: relation between the sickle cell gene and circulating plasma levels of testosterone, luteinising hormone (LH) and follicle stimulating hormone(FSH) in adult males. Clin Chim Acta. 1980;105:269–73.

23. Martins PRJ, De Vito FB, Resende GAD, Kerbauy J, Pereira G de A, Moraes-Souza H, et al. Male sickle cell patients, compensated transpubertal hypogonadism and normal final growth. Clin Endocrinol (Oxf). 2019;91(5):676–82.

24. Barbotin AL, Ballot C, Sigala J, Ramdane N, Duhamel A, Marcelli F, et al. The serum inhibin B concentration and reference ranges in normozoospermia. Eur J Endocrinol. 2015;172(6):669–76.

25. Kristensen DM, Desdoits-Lethimonier C, Mackey AL, Dalgaard MD, De Masi F, Munkbøl CH, et al. Ibuprofen alters human testicular physiology to produce a state of compensated hypogonadism. Proc Natl Acad Sci U S A. 2018;115(4):E715–24.

